# Assessing visible aerosol generation during vitrectomy in the era of Covid-19

**DOI:** 10.1101/2020.05.30.20117572

**Authors:** Sidath Liyanage, Pathma Ramasamy, Omar Elhaddad, Kieren Darcy, Andrew Hudson, Johannes Keller

## Abstract

**Objective:** To assess visible aerosol generation during simulated vitrectomy surgery. Methods: A model comprising a human cadaveric corneoscleral rim mounted on an artificial anterior chamber was used. Three-port 25 gauge vitrectomy simulated surgery was performed with any visible aerosol production recorded using high speed 4K camera. The following were assessed: (1) vitrector at maximum cut rate in static and dynamic conditions inside the model, (2) vitrector at air-fluid interface in physical model, (3) passive fluid-air exchange with a backflush hand piece, (4) valved cannulas under air, and (5) defective valved cannula under air.

**Results:** No visible aerosol or droplets were identified when the vitrector was used within the model. In the physical model, no visible aerosol or droplets were seen when the vitrector was engaged at the air-fluid interface. Droplets were produced from the opening of backflush hand piece during passive fluid-air exchange. No visible aerosol was produced from the intact valved cannulas under air pressure, but droplets were seen at the beginning of fluid-air exchange when the valved cannula was defective.

**Conclusions:** We found no evidence of visible aerosol generation during simulated vitrectomy surgery with competent valved cannulas. In the physical model, no visible aerosol was generated by the high-speed vitrector despite cutting at the air-fluid interface.

## Introduction

Surgical procedures involving the use of high-speed surgical devices such as drills are currently considered to be aerosol generating procedures (AGP). This aerosol is potentially able to carry the virus causing Covid-19 (1). There is debate within the Ophthalmology community regarding whether surgical procedures such as phacoemulsification and vitrectomy can be classified as AGPs and potentially transmit infective material (2,3). The purpose of this paper is to determine if visible aerosol may be generated in simulated vitrectomy surgery using high magnification and high speed video analysis in a human cadaveric model.

## Methods

This study adhered to the tenants of the Declaration of Helsinki. A fresh human cadaveric corneoscleral rim was obtained from the NHS Blood and Transplant for research purposes. The rim was fitted on a Coronet™ artificial anterior chamber (AAC) (Network Medical, Ripon, UK) and was mounted within a model head used for cataract training (Phillips Studio, Bristol, UK). A Constellation vitreoretinal surgery platform (Alcon, Camberley, United Kingdom) was set up for vitrectomy surgery. Three 25-gauge valved 4mm cannulas were placed 2mm behind the limbus, just anterior to the junction of the scleral rim and AAC. Aerosol generation was assessed using high speed 4K camera (Canon 5D mark IV, 100mm prime macro lens; Canon, Tokyo, Japan, with a Hoya Pro1 Digital +3 Close-up lens; Kenko Tokina Co., Tokyo, Japan) mounted on a tripod and fixed lighting sources (Rotolight Neo II; Rotolight, Iver Heath, UK). Vitrectomy was performed at 7500 cuts per minute (cpm), a vacuum of 550mmHg and an infusion pressure of 30mmHg under the following experimental settings:

### A. Vitrectomy in static positions

The port was placed in the centre of the model, 1mm past the internal os of the 4mm valved cannula (shaft of vitrector marked at 5mm from port) and within the valved cannula.

### B. Dynamic vitrectomy

Using the same parameters, the experiment was repeated by advancing the vitrector further into the chamber accompanied by dynamic movements to simulate vitrectomy surgery.

### C. Vitrector in fluorescein solution

Fluorescein 2% (Minims; Bausch & Lomb, Kingston, United Kingdom) was diluted with balanced salt solution (BSS; Balanced salt solution; Beaver-Visitec International, Oxford, UK) to 1% concentration and placed in a shallow container. Ultraviolet illumination (Kam UV bar, Dunstable, United Kingdom) was used to excite the fluorescein. The vitrector was repeatedly moved back and forth through the room air-liquid fluorescein interface (4).

### D. Passive extrusion during fluid-air exchange

A backflush cannula (Surgistar, California, USA) was introduced into the fluid-filled chamber and air infusion pressure set at 45mmHg to perform passive fluid-air exchange (FAX).

### E. Air infusion with intact cannula valve

Instruments were withdrawn from the model and air infusion pressure set at 45mmHg with intact cannula valves.

### F. Air infusion with damaged cannula valve

The vitrector was used to damage a valved cannula in the fluid-filled model. Air infusion pressure was set at 45mmHg with passive FAX occurring through the leaking cannula valve.

## Results

We found that no visible aerosol was noted outside the model when the high-speed vitrector was engaged inside the model at 7500 cpm, despite varying the location of the vitrector probe while cutting and making exaggerated manoeuvres within the model. High magnification and high speed video analysis of the external os of the valved cannula did not identify any aerosol produced. In the simulation using diluted fluorescein, no aerosol was noted when repeatedly breaching the air-fluid interface with the vitrector engaged. No aerosol was identified originating from the competent valved cannula during FAX. When using a backflush hand piece to achieve passive extrusion during FAX, large droplets and no aerosol were seen egressing from the vent on the hand piece. When the valve in the cannula was damaged, droplets were noted at the external os of the cannula at the beginning of FAX.

## Discussion

We sought to to determine if visible aerosol is generated in simulated vitrectomy surgery. Other studies published online have analysed vitrectomy in plastic eye models and found no aerosol generation (4). Our study used human cadaveric corneoscleral tissue mounted on an artificial anterior chamber to more accurately simulate the biomechanics of human tissue and high-speed videography to highlight any visible aerosol generation during vitrectomy surgery. While the cannulas were sited closer to the limbus in this model than in standard clinical practice, it is unlikely that this difference may influence aerosol generation.

In this context, aerosol generation requires enough energy (usually mechanical or airflow) directed at a gas-fluid interface to disrupt the surface tension and allow aerosol particles to form. Following this, airflow is required to entrain these particles (5). High magnification and slow-motion analysis of the vitrector at 7500 cpm did not reveal any aerosol production at the external os of the cannula when the vitrector is inside the model or at the air-fluorescein interface. The latter physical model raises three possibilities: (1) the high-frequency back-and-forth motion of the guillotine blade does not dispense enough energy to cause visible aerosol, or (2) the direction or diffusion of energy release may not disrupt the interface sufficiently, or (3) any droplets or aerosol formed by the blade at the interface are immediately aspirated by the vacuum required for vitrectomy.

The intraocular environment is less likely to favour aerosol generation. In the majority of cases vitrectomy is conducted without air-fluid interface. In the absence of an air-fluid interface aerosol production is impossible. Some surgeons advocate interface vitrectomy for complex detachment surgery (6). In these situations, if the energy of the vitrector is sufficient to produce aerosol and this aerosol escapes the vacuum, its egress from the eye would be prevented by the valved cannulas. The trapped aerosol would either settle on the intraocular surfaces or merge with the fluid interface. The only potential site for aerosol production while the vitrector is in situ is at the mouth of the valved cannula. However, there is no high speed movements or mechanical energy exerted at this point making aerosol production unlikely, as evidenced by the video which shows a droplet of fluid slowly emerging.

Another possible cause of aerosol generation during vitrectomy is FAX where pressurised air is used to replace the fluid in the vitreous cavity. At the beginning of FAX there is no distinct interface present as air is directly introduced into the fluid. This creates bubbles that coalesce due to Laplace pressure, creating an air-fluid interface. As more fluid is drained out, there is a potential for aerosol generation as the stream of air may disrupt the surface during extrusion. However, any aerosol produced would be trapped by the valves and either settle on the intraocular surfaces or rejoin the fluid meniscus.

The only risk of escape to the atmosphere is during passive extrusion if the cannula hovers above the air-fluid interface as aerosol is being created. This may be vented through the cannula and backflush hand piece. If this occurs, some aerosol would settle on the internal wall of the cannula or hand piece before being vented. No aerosol was noted during passive extrusion in the model. This situation may be avoided with using active extrusion for FAX. Another possibility for aerosol generation depends on the failure of the valved cannula. In this scenario, the high flow of air can cause a stream of droplets to be forced through the cannula at the beginning of FAX as demonstrated in this study. This is unlikely in clinical practice as FAX is usually commenced with either an extrusion cannula or vitrector in situ.

This experiment utilised an optical setup with an effective pixel size of 3 μm (5.36 μm pixel size of camera coupled with 1.75× optical magnification) and identified aerosol during active phacoemulsification (7). It allows accurate visualisation of 6 μm and larger particles every 30 ms. It is possible to discern particles smaller than this threshold due to light diffraction if their density is high. This threshold is lowered when using fluorescein and ultraviolet excitation for added contrast. Due to this, we are unable to comment on whether aerosol particles too small to be visualized at low density were created or whether such particles could be produced in the absence of visible aerosol. These particles may remain suspended for a long time if they are generated in a closed room with no airflow. In an operating theatre, the concentration of these airborne particles would be diluted by the positive air pressure generating at least 20 air changes per hour as specified in the Royal College of Ophthalmologists specifications for these facilities (8). Further, the downward flow patterns generated by standard ventilation systems will encourage these particle to settle on surfaces (9). This emphasises the importance of meticulous hand hygiene.

Currently, the presence of SARS-CoV-2 in intraocular fluid is unknown. Studies to determine the presence of SARS-CoV-2 in intraocular fluids would improve our understanding of the risk of transmission during intraocular surgery. If SARS-CoV-2 particles are present in intraocular fluid, any viral load is likely to be significantly and serially diluted by the infusion of balanced salt solution that is necessary for vitrectomy. This follows the principles of first order decay and is dependent on the inflow rate of BSS infusion, the duration of vitrectomy and the outflow of BSS through aspiration. Consequently, it is uncertain whether any aerosol generated would pose a risk of transmitting Covid-19 disease to healthcare workers by containing enough viable SARS-CoV-2 to be an effective inoculum. This study did not identify any visible aerosol generation in simulated vitrectomy in a human cadaveric model.

## Data Availability

All data included in manuscript.

## Acknowledgements

We wish to acknowledge the support of NHS Blood and Transplant.

## Funding

The authors received no financial support for the research, authorship, and/or publication of this article.

**Figure 1:**
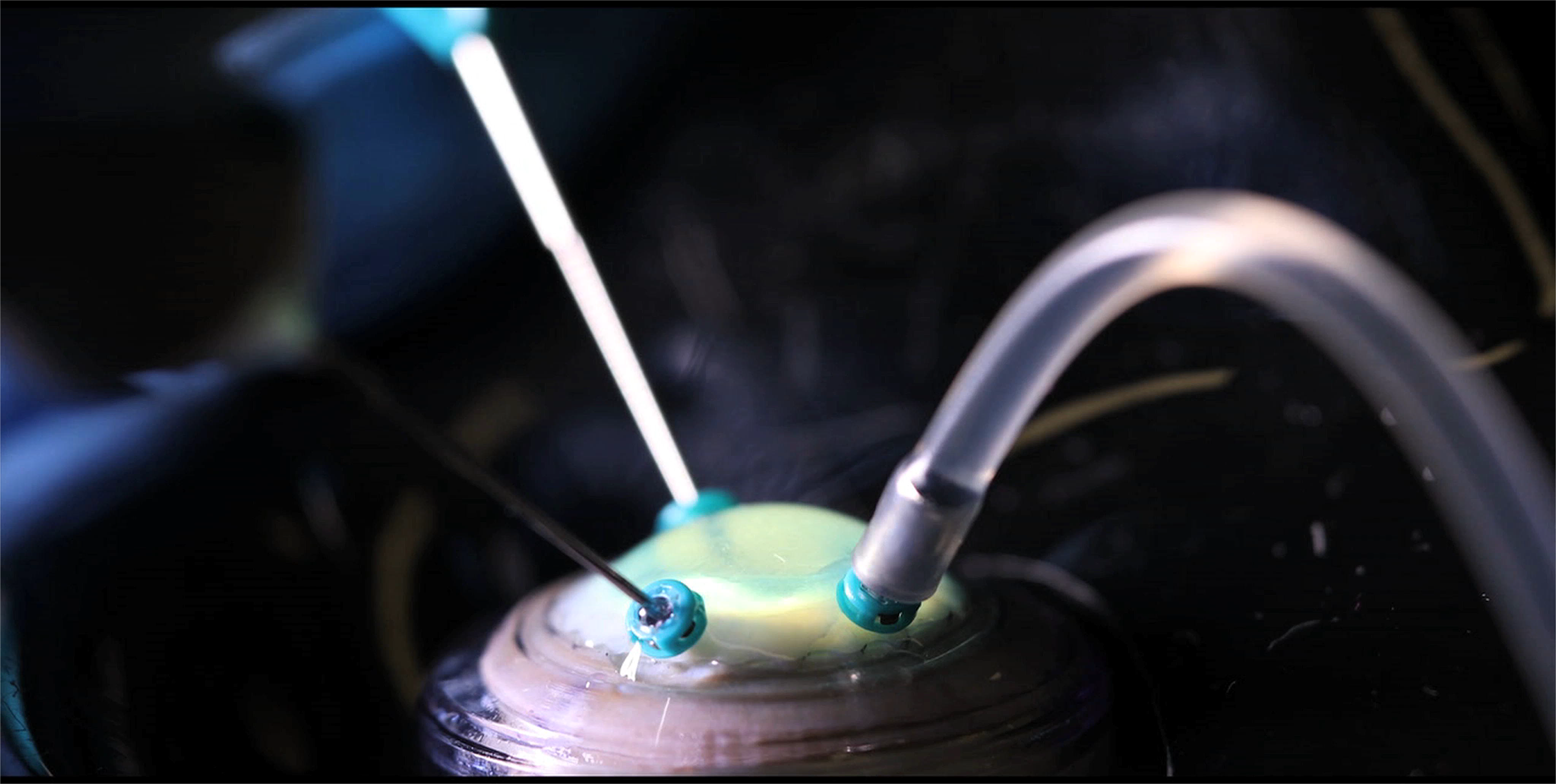
Photograph showing the corneoscleral rim mounted on the anterior artificial chamber. This is sited within a model head used for cataract training. A standard 25g 3-port pars plana vitrectomy set up is used.

**Figure 2:**
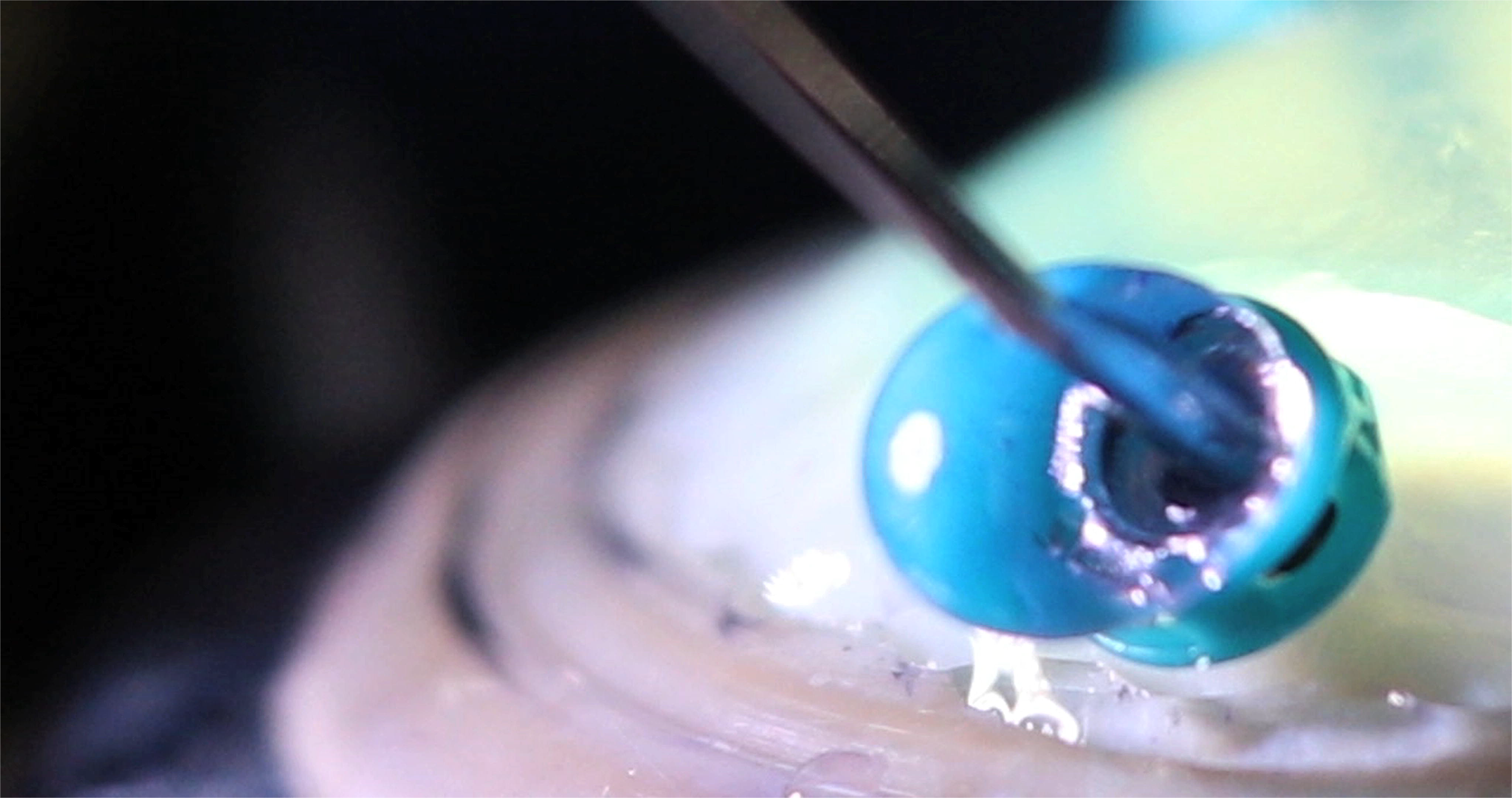
Photograph showing droplet formation but no visible aerosol at the external os of the valved cannula during vitrectomy at 7500 cpm.

**Figure 3:**
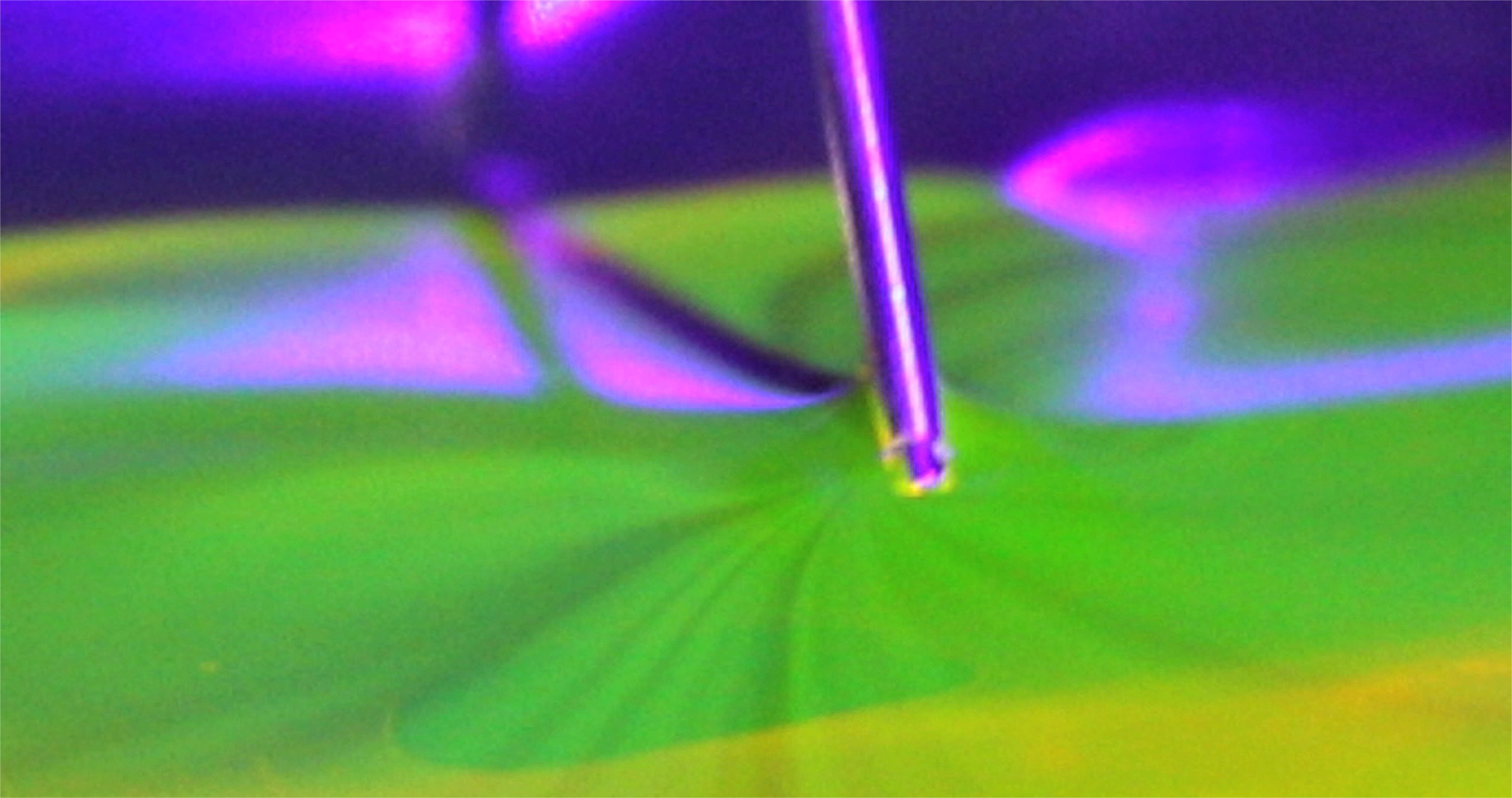
Photograph showing no visible aerosol production when the vitrector is engaged at the air-fluorescein interface as visualised with ultraviolet light

**Figure 4:**
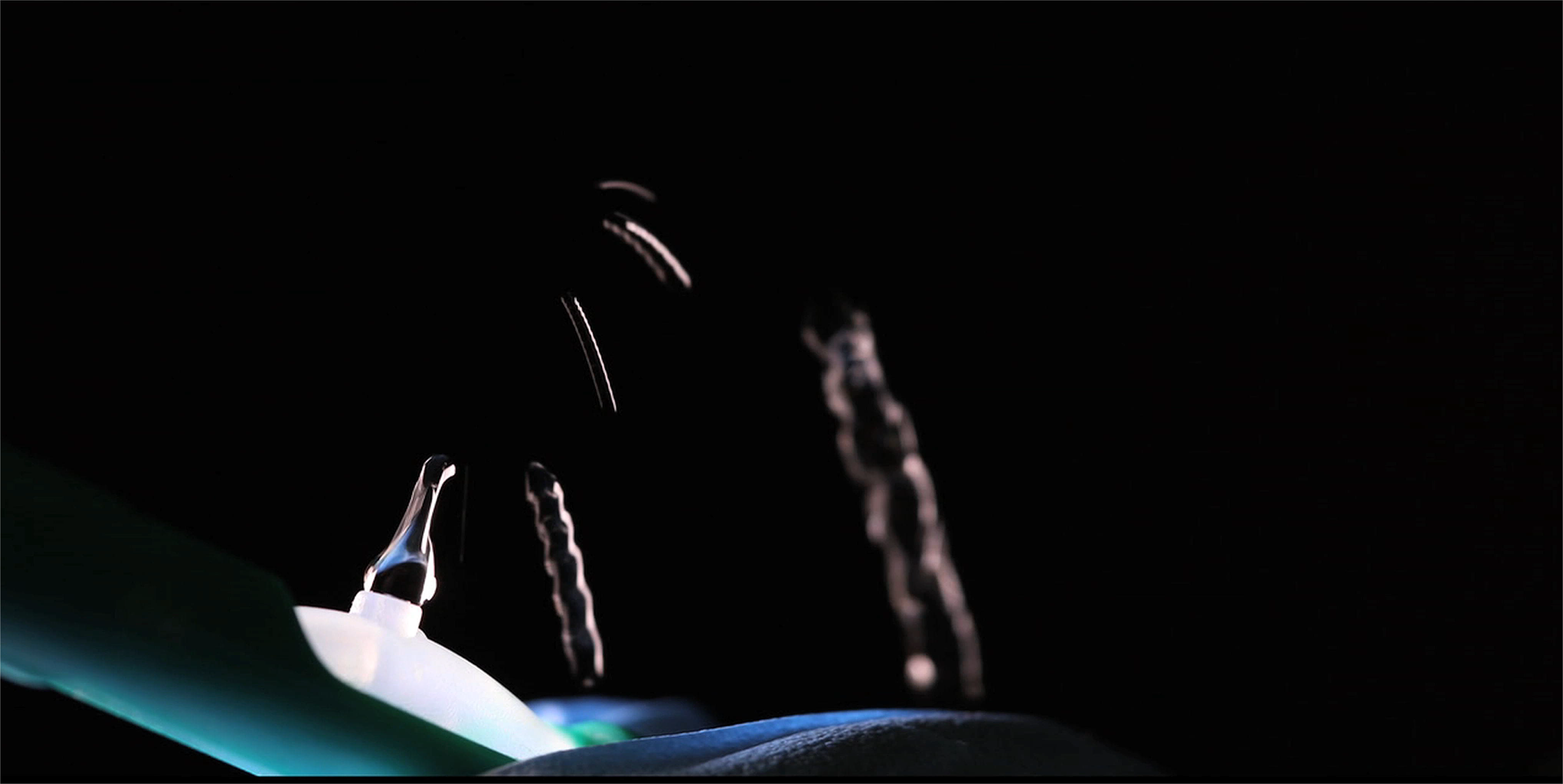
Photograph of droplets of fluid being vented during passive extrusion at fluid-air exchange

**Figure 5:**
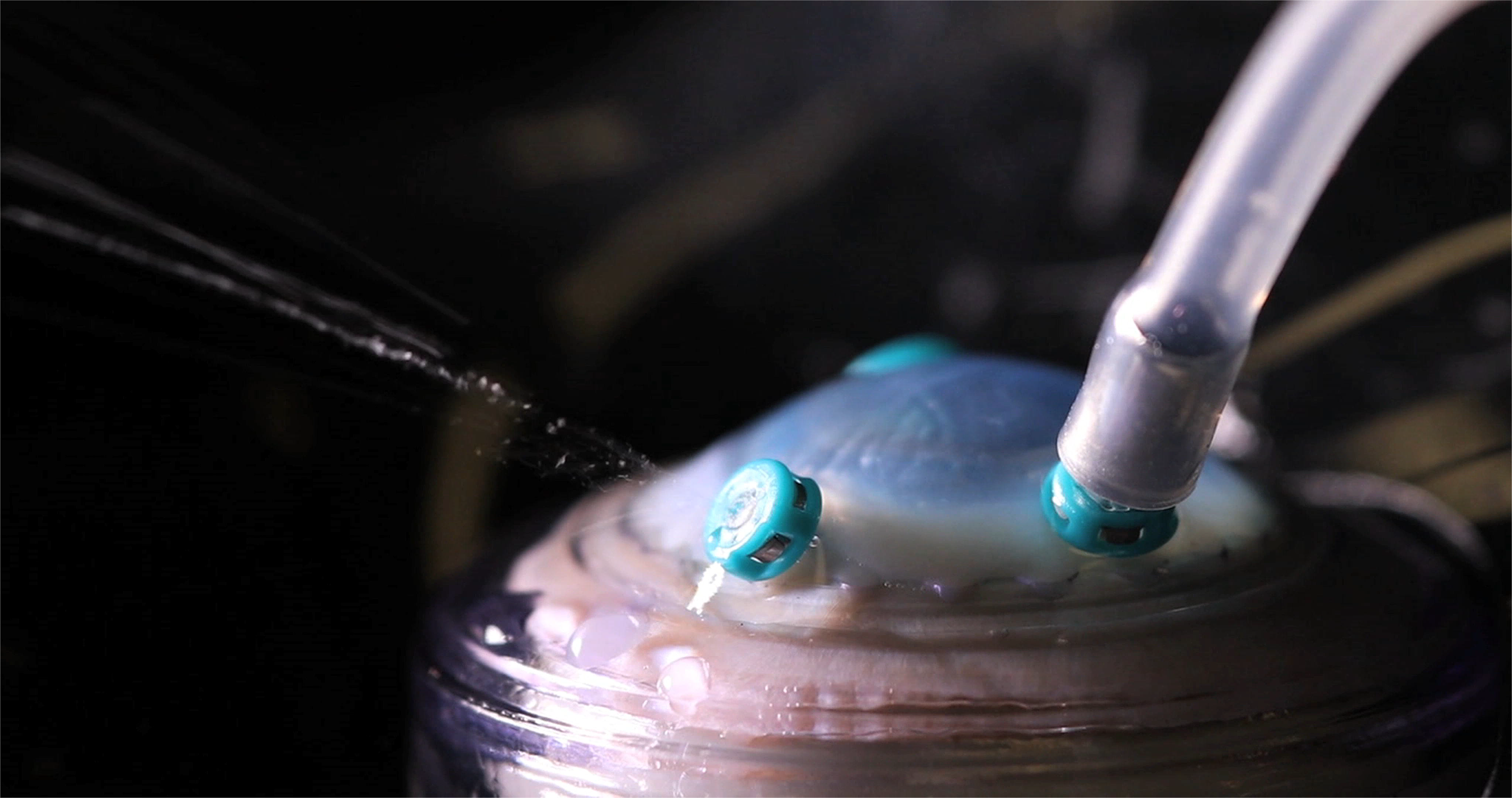
Photograph of droplets vented from a failed valved cannula at the beginning of fluid-air exchange

